# Health concerns and government distrust: Variation in types of COVID-19 vaccine hesitancy by racial and ethnic group before and at universal vaccine eligibility in the US

**DOI:** 10.1101/2025.07.04.25330884

**Authors:** Meg Ellingwood, Alex Reinhart, D. Phuong Do, Robin Mejia

**Affiliations:** Dept. of Statistics & Data Science, Carnegie Mellon University, Pittsburgh, PA, USA; Public Health Policy, Zilber School of Public Health, University of Wisconsin–Milwaukee, Milwaukee, WI, USA; Dept. of Biostatistics, School of Public Health, University of Washington, Seattle, WA, USA

## Abstract

The development and deployment of COVID-19 vaccines were crucial to combating the global pandemic, but vaccine hesitancy posed a challenge and remains a crucial problem today. Understanding why people delay or refuse vaccination can help with current and future vaccine campaigns by suggesting both target audiences and messaging. Because hesitant individuals report different reasons for their hesitancy, distinguishing between different types of hesitancy can help identify who will benefit from which strategies. Using 311,494 responses from the U.S. COVID-19 Trends and Impact Survey (CTIS), we used Latent Class Analysis to examine distributions of concurrently reported reasons for hesitancy using latent class analysis, comparing distributions of responses across racial and ethnic groups. Using responses from both the early phase of vaccine availability and after all U.S. adults became eligible in the spring of 2021, we investigated differences between prospective vs. lived hesitancy. LCA revealed three different types of COVID-19 vaccine hesitancy. Some respondents primarily reported health-related concerns as reasons for hesitancy, while others indicates distrust of the government and vaccines, but the majority report fewer distinct reasons. In February 2021, White, Black, and Hispanic respondents had similar distributions of these types, but by May the results diverged. In May, White respondents were more likely to report trust concerns than Black or Hispanic respondents, who were more likely to report health concerns as their reasons for continued hesitancy. Our results contribute to the development of a more nuanced picture of COVID-19 vaccine hesitancy and the motivations behind it. The distinction between *health-concerned* and *distrustful* types of vaccine hesitancy highlights the importance of confidence in vaccine uptake, and in targeted strategies to address address hesitancy.

## Introduction

The development and delivery of effective COVID-19 vaccines was a milestone public health achievement, saving lives and enabling a return to normal life. In December 2020, nearly a year after the first COVID-19 case was diagnosed, the first two COVID-19 vaccines received emergency use authorization in the United States. The Centers for Disease Control and Prevention (CDC) offered guidelines for phased distribution of the initially limited supply, and states implemented a range of distribution policies. Broadly, healthcare workers and residents of long-term care facilities were the first to be eligible for the vaccine, followed by everyone over the age of 75. Access expanded over the following weeks, with specifics decided by each state. Finally, by May 2021, all adults were eligible [1, 2].

Vaccine hesitancy, already concern in public health, was anticipated to be a problem with the COVID-19 vaccine [3]. Traditional frameworks for vaccine hesitancy highlight the importance of convenience, confidence, and complacency as factors behind vaccine hesitancy or demand [4]. However, the COVID-19 pandemic created new context [5, 6]. The extreme salience of the disease was predicted to motivate uptake [7]. However, some Americans showed concerns about the development and approval process [8]. Additionally, because the vaccine was authorized in December 2020 and many people did not have access until late spring 2021, initial studies assessed how people believed they would respond when a vaccine became available, rather than how they actually responded, often much later.

Previous work both before and during the vaccine rollout identified many individual factors associated with hesitancy toward the COVID-19 vaccine, including gender, age, income, education, and occupation [8–17].

Hesitant individuals reported many reasons, from worries about safety and side effects to distrust of the government and the development process for the new vaccines [9, 15].

Race and ethnicity were also associated with hesitancy in the U.S. [9, 16], with vaccine hesitancy higher among many non-White groups early on, with the exception of Asians. This was particularly concerning given the disproportionate impact of the COVID-19 pandemic in Black and Hispanic communities.

The reasons given for hesitancy in Spring 2021 also varied among different demographic groups. Overall, fear of side effects was the single most commonly reported reason within each racial/ethnic group, but the next-most common reason differed by race/ethnicity: White respondents were more likely to say they didn’t trust the government compared with wanting to wait and see if the vaccine was safe, while the reverse was true of Hispanic and Black respondents [12].

In survey research on COVID-19 vaccine hesitancy, numerous reasons for hesitancy were potentially interconnected. However, studies commonly analyzed reasons selected for vaccine hesitancy individually, ignoring relationships between reasons. Further, most studies snapshots in time. In this study of COVID-19 vaccine hesitancy, we employ Latent Class Analysis to identify groups of hesitancy reasons that tend to be reported in conjunction and model those relationships to identify types of hesitancy. We then examine how the types of hesitancy vary between different racial/ethnic groups and across time.

## Methods

### Data Summary

The US COVID-19 Trends & Impact Survey (CTIS) was a large, daily cross-sectional online survey conducted by the Delphi Group at Carnegie Mellon University in partnership with Meta and the University of Maryland from April 2020 through June 2022 [18]. Participants were recruited from the Facebook monthly adult (age 18+) active user base using stratified random sampling via an invitation above their News Feed. Those who responded were sent to a Qualtrics survey administered by Carnegie Mellon University. Survey respondents gave written informed consent to participate. The survey did not collect any personally identifying information [18, 19].

Questions on vaccine hesitancy were introduced on February 8, 2021, and remained unchanged through May 20, 2021. Our analytical samples are composed of respondents from February 8 through February 28, during a period when vaccines were only available to a limited subset of the population (*prospective hesitancy* sample) and from May 1 to May 20, when all adults were eligible for the vaccine (*lived hesitancy* sample). Data for this study were accessed April 18, 2022.

For full questions and answer choices used in this analysis are provided, see S1 Appendix. Respondents who selected that they had not yet received one or more doses of a COVID-19 vaccine, and responded that they “probably would not” or “definitely would not” accept a vaccine if one were offered to them that day, were classified as vaccine hesitant. Hesitant respondents received a follow-up question asking them to select one or more reasons for choosing not to be vaccinated from a list of 15 options: concerns about side effects, having an allergic reaction, unsure if the vaccine will work, don’t believe they need it, don’t like vaccines generally, doctor has not recommended, want to wait and see if it is safe, others need it more, worried about cost, don’t trust the government, against religious beliefs, don’t trust COVID-19 vaccines specifically, health condition, pregnant/breastfeeding, and other.

Race and ethnicity of respondents were ascertained through a two-part question that asked about Hispanic or Latino origin and race. Race-stratified analyses were conducted with respondents who identified as non-Hispanic White, non-Hispanic Black, or Hispanic, as those groups had adequate sample size for analysis.

### Latent Class Analysis

To assess how respondents tended to select multiple hesitancy reasons together, we performed latent class analysis (LCA), a form of mixture model in which individuals are assumed to belong to unobserved groups or “latent classes” [20]. These unobserved latent classes are modeled based on observed categorical variables. In this study, we applied LCA to assess patterns for COVID-19 vaccine hesitancy by modeling latent classes of “hesitancy types”, based on reasons selected by survey respondents. Each latent class represents a distinct response pattern, with a vector indicating the probabilities of selecting specific hesitancy reasons. For example, a respondent in Hesitancy Class A has a certain probability of selecting Hesitancy Reason X. This approach enables the estimation of common hesitancy response patterns and the probability that each respondent belongs to each class. We included race/ethnicity as a categorical grouping variable, allowing different latent structures across race/ethnicity [21]. This enabled comparison of the identified classes, and the response probabilities, between racial/ethnic groups.

Goodness-of-fit tests using likelihood ratios and AIC were used to compare possible models [22]. The final number of latent classes selected was based on balancing the results of the likelihood ratio tests and AIC with interpretability of the resulting model. All analyses were conducted in R (Version 4.1.1, [23]), using the glca package [24].

### Sensitivity Analysis

Because incorporating response weights was not feasible for the modeling methods used in this study, all results reported are based on unweighted survey data. As a sensitivity analysis, we compared the distribution of survey weights by class membership probabilities, to assess whether the size of classes would have been affected by inclusion of survey weights.

### Ethics Statement

Survey respondents gave written informed consent to participate in CTIS. This analysis was allowed under the overall survey consent. The Carnegie Mellon University Institutional Review Board approved the study (STUDY2022 00000113).

## Results

### Study Sample and Participant Characteristics

The sample selection is shown in Fig 1. To evaluate prospective and lived hesitancy, we analyzed survey responses from February (N = 857,563) and May 2021 (N = 562,572) separately. Because previous analysis of CTIS data (e.g., [16]) identified high rates of adversarial response patterns among respondents who selected self-described gender (e.g., writing in anti-trans slurs), we excluded participants who selected self-described gender from the analysis (N = 4,766, 0.55%, in February; N = 4,094, 0.73%, in May). We then identified respondents who were unvaccinated. From those, we selected those who indicated they would “probably not” or “definitely not” accept a vaccine if they were offered it on that day (N = 236,677 in February and N = 74,817 in May). We then excluded participants who did not respond to the question asking the reasons for their hesitancy, yielding an analytical study sample of 159,755 individuals in February, and 73,362 individuals in May.

**Fig 1.**
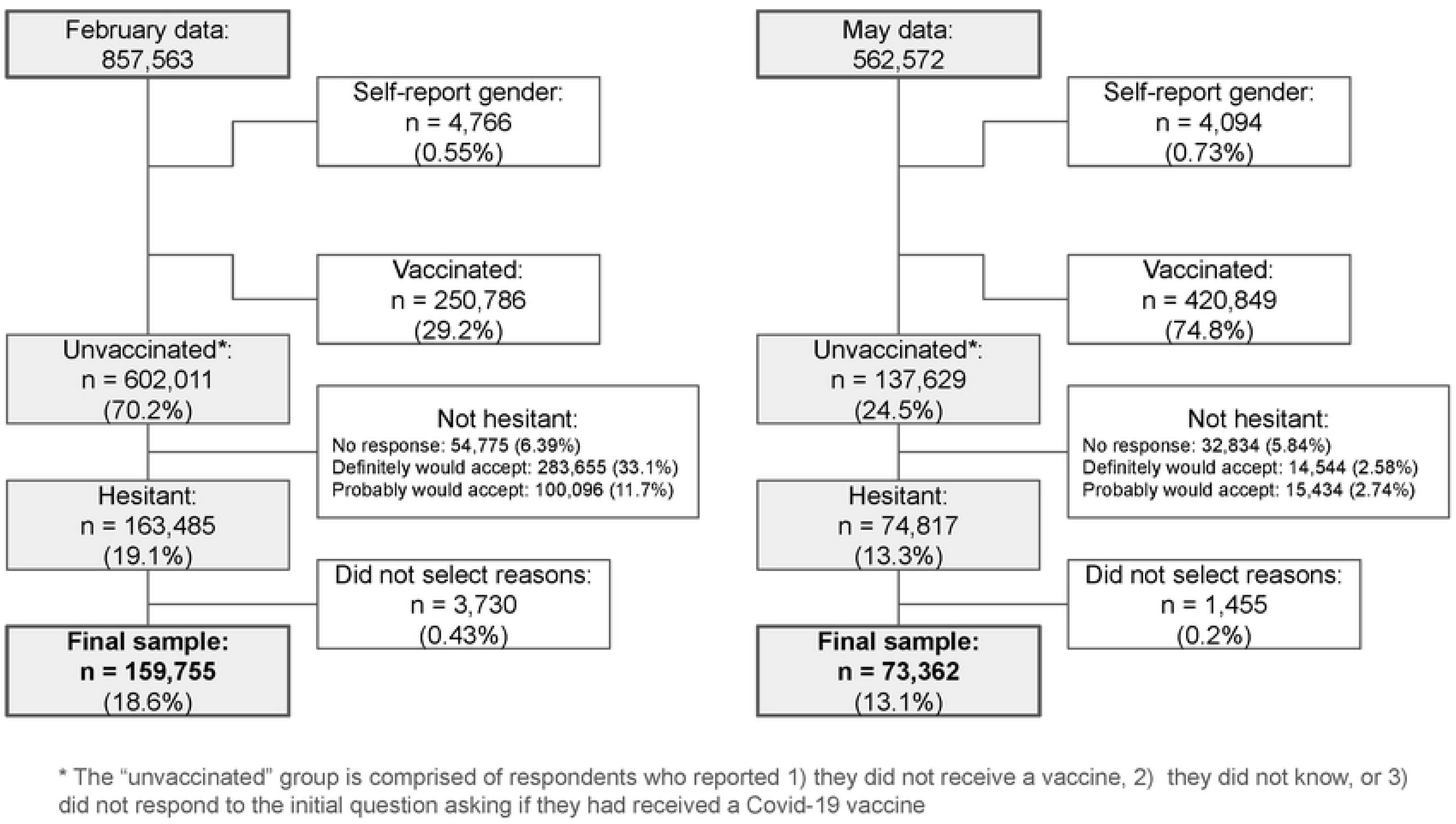
Processing of respondents to isolate “hesitant” sample. Latent class analysis was performed only on participants who responded to the item asking their reasons for hesitancy. Percentages at each stage are out of the original sample size (i.e., all February or all May respondents).

The demographic makeup of the February and May samples is presented in Table 1. Demographic patterns were broadly similar across the two time periods. The sampling process permitted respondents from February to be sampled in May, and privacy considerations in the survey design prevented tracking of respondents over time; however, any overlap in the samples is expected to be small.

**Table 1.**
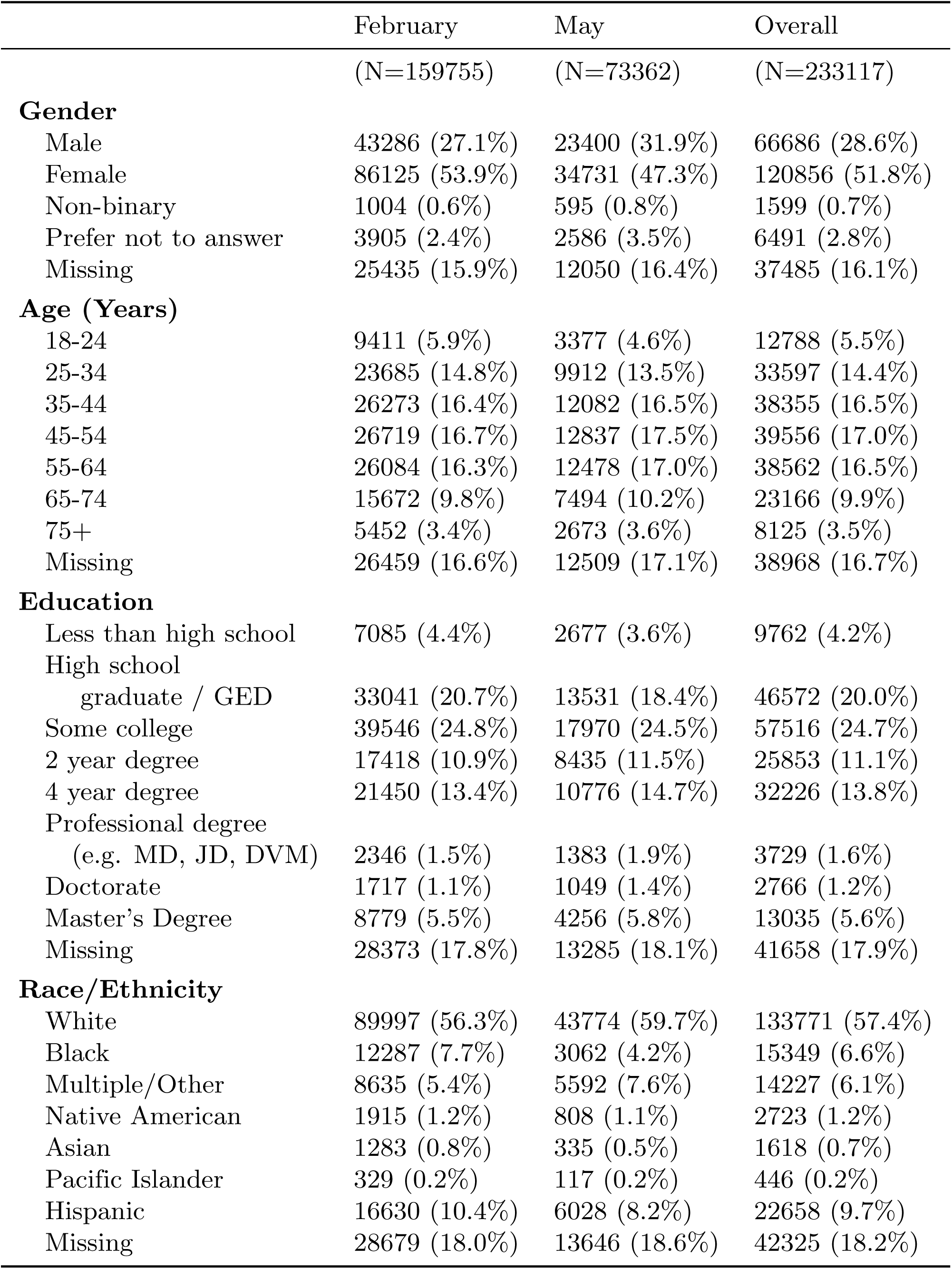
Demographics of final sample of hesitant participants who reported their hesitancy reasons.

|  | February<br>(N=159755) | May<br>(N=73362) | Overall<br>(N=233117) |
| --- | --- | --- | --- |
| **Gender** |  |  |  |
| Male | 43286 (27.1%) | 23400 (31.9%) | 66686 (28.6%) |
| Female | 86125 (53.9%) | 34731 (47.3%) | 120856 (51.8%) |
| Non-binary | 1004 (0.6%) | 595 (0.8%) | 1599 (0.7%) |
| Prefer not to answer | 3905 (2.4%) | 2586 (3.5%) | 6491 (2.8%) |
| Missing | 25435 (15.9%) | 12050 (16.4%) | 37485 (16.1%) |
| **Age (Years)** |  |  |  |
| 18-24 | 9411 (5.9%) | 3377 (4.6%) | 12788 (5.5%) |
| 25-34 | 23685 (14.8%) | 9912 (13.5%) | 33597 (14.4%) |
| 35-44 | 26273 (16.4%) | 12082 (16.5%) | 38355 (16.5%) |
| 45-54 | 26719 (16.7%) | 12837 (17.5%) | 39556 (17.0%) |
| 55-64 | 26084 (16.3%) | 12478 (17.0%) | 38562 (16.5%) |
| 65-74 | 15672 (9.8%) | 7494 (10.2%) | 23166 (9.9%) |
| 75+ | 5452 (3.4%) | 2673 (3.6%) | 8125 (3.5%) |
| Missing | 26459 (16.6%) | 12509 (17.1%) | 38968 (16.7%) |
| **Education** |  |  |  |
| Less than high school | 7085 (4.4%) | 2677 (3.6%) | 9762 (4.2%) |
| High school graduate / GED | 33041 (20.7%) | 13531 (18.4%) | 46572 (20.0%) |
| Some college | 39546 (24.8%) | 17970 (24.5%) | 57516 (24.7%) |
| 2 year degree | 17418 (10.9%) | 8435 (11.5%) | 25853 (11.1%) |
| 4 year degree | 21450 (13.4%) | 10776 (14.7%) | 32226 (13.8%) |
| Professional degree<br>(e.g. MD, JD, DVM) | 2346 (1.5%) | 1383 (1.9%) | 3729 (1.6%) |
| Doctorate | 1717 (1.1%) | 1049 (1.4%) | 2766 (1.2%) |
| Master's Degree | 8779 (5.5%) | 4256 (5.8%) | 13035 (5.6%) |
| Missing | 28373 (17.8%) | 13285 (18.1%) | 41658 (17.9%) |
| **Race/Ethnicity** |  |  |  |
| White | 89997 (56.3%) | 43774 (59.7%) | 133771 (57.4%) |
| Black | 12287 (7.7%) | 3062 (4.2%) | 15349 (6.6%) |
| Multiple/Other | 8635 (5.4%) | 5592 (7.6%) | 14227 (6.1%) |
| Native American | 1915 (1.2%) | 808 (1.1%) | 2723 (1.2%) |
| Asian | 1283 (0.8%) | 335 (0.5%) | 1618 (0.7%) |
| Pacific Islander | 329 (0.2%) | 117 (0.2%) | 446 (0.2%) |
| Hispanic | 16630 (10.4%) | 6028 (8.2%) | 22658 (9.7%) |
| Missing | 28679 (18.0%) | 13646 (18.6%) | 42325 (18.2%) |

### Classes of Prospective Hesitancy, February 2021

In February, 19.06% of all respondents, and 27.2% of unvaccinated respondents, were identified as hesitant. Because the COVID-19 vaccine was not yet widely available during this time period, we consider this hesitancy to be primarily prospective: people’s belief about what they would do when a not-yet-possible option became available.

We initially fit latent class models with 3 to 30 classes. As the number of classes increased, AIC consistently decreased. Likelihood ratio tests confirmed improved model fit with more classes, likely because of the large sample size. Because interpretability becomes increasingly difficult as the number of classes increases, we restricted further analysis to models with fewer than 10 classes. Examination of these models revealed that beyond three classes, allowing additional classes primarily subdivided already small groups without providing meaningful distinctions. Thus, for parsimony, we determined that three classes provided the best balance between model fit and interpretability.

### Among All Race/Ethnicity Groups

The largest class had an average membership probability of 55.1% (Fig 2, panel A; S1 Appendix provides point estimates and standard errors). On average, respondents in this class selected 1.61 reasons, indicating a tendency to choose one to two specific reasons for hesitancy. No particular reasons stood out for this group. Wanting to “wait and see” if the vaccines were safe had a probability of 25.8% to be selected, but several other reasons had comparable chance of selection, including that they “don’t trust COVID-19 vaccines” (19.5%). In contrast, “don’t like vaccines” in general had a much lower probability of only 6.2%. We labeled this group the *ambiguous* class.

**Fig 2.**
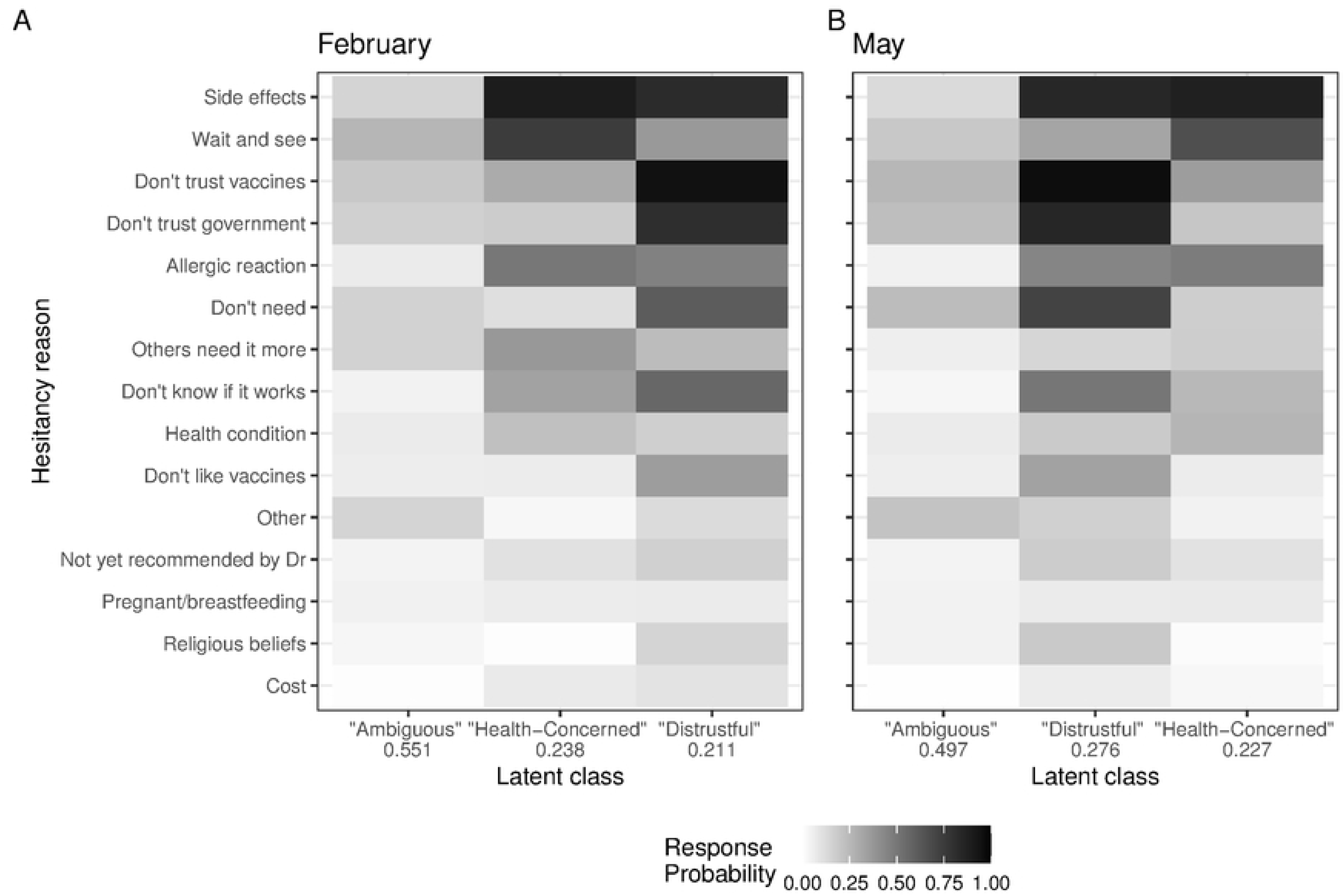
Latent class analysis among all race/ethnicity groups. A: Results of LCA with three classes on full sample of participants in February 2021. Hesitancy reasons are arranged by overall descending prevalence (regardless of class) in the February sample. B: Results of LCA with three classes on full sample of participants in May 2021.

The second class had an average membership probability of 23.8%. Unlike the first class, multiple reasons emerged as notable. The top two reasons were “concern about possible side effects”, with a response probability of 89.8%, and the desire to “wait and see” if the vaccine was safe, with a probability of 74.7%. Members of this class were also more likely than other classes to express concerns about “allergic reactions” and feelings that “others need it more”. Some also indicated they “don’t know” if the vaccine would work. The average number of reasons selected by this class was 4.01. Since the most prominent reasons for this class were related to worries about well-being and the effects of the vaccine on their immediate health, we labeled this group the *health-concerned* class.

The third class was slightly smaller, with an average membership probability of 21.1%. However, it had the highest average number of selected reasons, at 5.96. The most commonly selected reason was that they did not trust the COVID-19 vaccines, with a probability of 94.8%. Respondents in this class also frequently indicated that they were worried about side effects (82.6%) and that they did not trust the government (80.9%). They were more likely than those in other classes to report that they “don’t like vaccines” in general, and also that they “do not need” the vaccine. Because distrust-related reasons were central to this class, we labeled it the *distrustful* class.

### Across Race/Ethnicity

Three-class LCA models fitted separately for the White, Black, and Hispanic respondents revealed that within each racial/ethnic group, an ambiguous class, a health-concerned class, and a distrustful class consistently emerged. The classes also exhibited similar relative proportions to each other: the ambiguous class was the largest, followed by health-concerned, and then distrustful. Response probabilities and relative class membership probabilities for each group’s model are displayed in Fig 3; probabilities on the scale of 0 to 1 with standard errors are provided in S1 Appendix.

**Fig 3.**
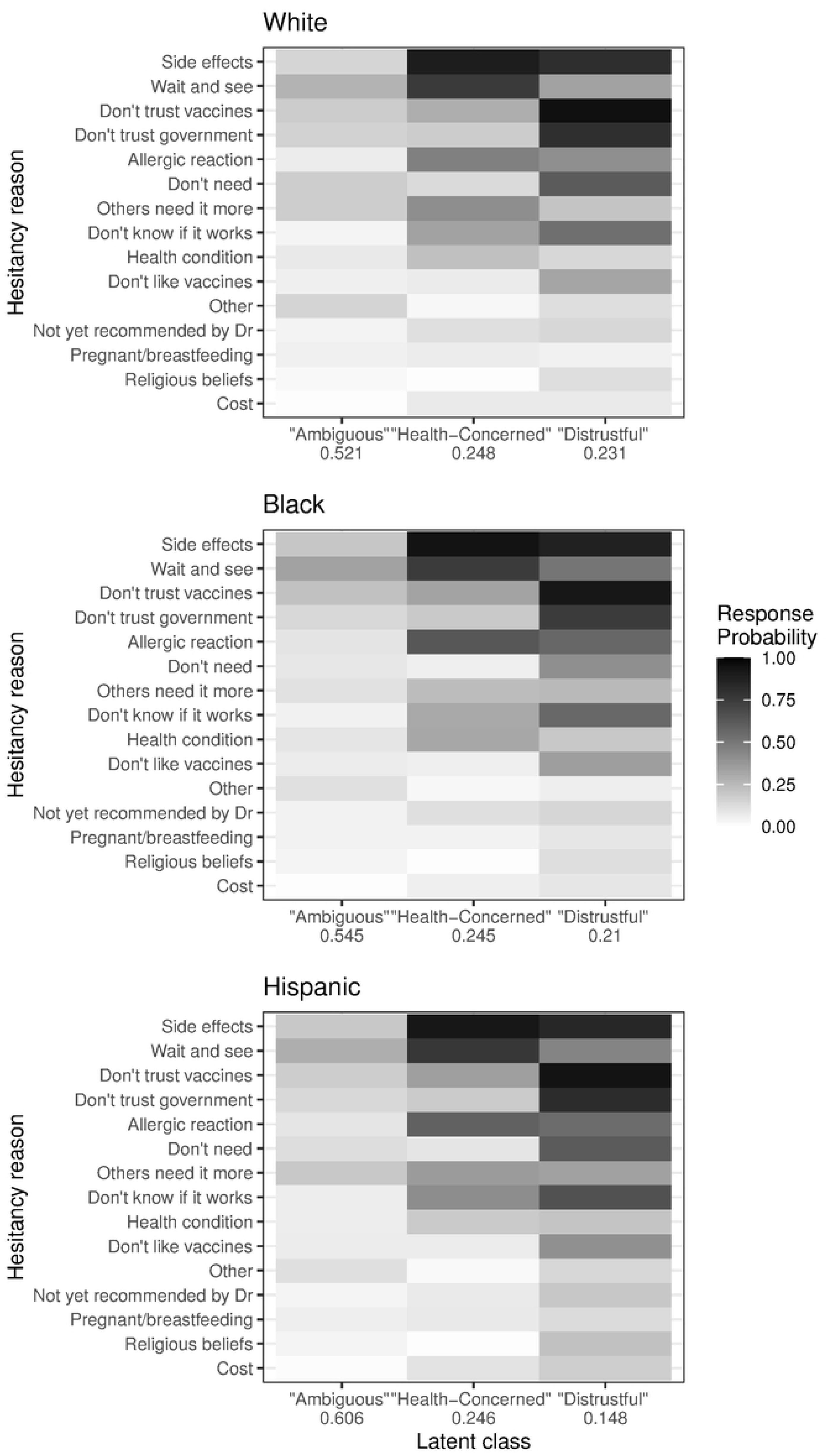
Results of LCA with three classes for White, Black, and Hispanic respondents in February 2021. Hesitancy reasons are arranged according to overall prevalence (regardless of class or race/ethnicity) in the February sample.

However, there were subtle differences in the distribution of reasons within each class across racial/ethnic groups. For example, in the “distrustful” class, Black respondents were 15 percentage points more likely to report concerns about “allergic reactions” than their White counterparts (see S1 Appendix). They were also 17 percentage points more inclined to indicate that they wanted to “wait and see” if the vaccine was safe. Similarly, Hispanic respondents were more likely to select health-related reasons within the “distrustful” class, compared to “distrustful” White respondents.

### Three Types of Lived Hesitancy, May 2021

In May, 13.3% of all respondents and 54.4% of respondents who were unvaccinated were coded as vaccine hesitant. As the vaccine was available to all US adults at this point, we consider this to be lived vaccine hesitancy. Using the sample that provided reasons for their hesitancy, we again fit models with 3 to 30 classes and observed that AIC improved with additional classes. Again, the addition of classes resulted in splitting already-small classes, and for parsimony and facilitation of comparison across time, we opt again to proceed with three classes.

### Among All Race/Ethnicity Groups

Response probabilities and relative class membership probabilities for May 2021 are displayed in Fig 2, panel B (S1 Appendix provides point estimates with standard errors.) The three-class LCA model fit on the May 2021 sample resulted in similar classes of response patterns found for the February prospective hesitancy sample. Characterized by relatively low probability of selection across many reasons, the ambiguous class was once again the largest. Individuals in this class selected an average of 1.66 reasons, and the probability of class membership was 49.7%, slightly lower than in February. However, by May 2021, the relative prevalence of the two remaining classes had flipped. The second-largest class was the distrustful class, with a membership probability of 27.6%, and the health-concerned class was the smallest, with a membership probability of 22.7%.

### Across Race/Ethnicity

Consistent with the February analysis, we again fit three-class LCA models for the May 2021 White, Black, and Hispanic respondents separately. The response probabilities and class membership probabilities for these models are presented in Fig 4; S1 Appendix provides point estimates and standard errors.

**Fig 4.**
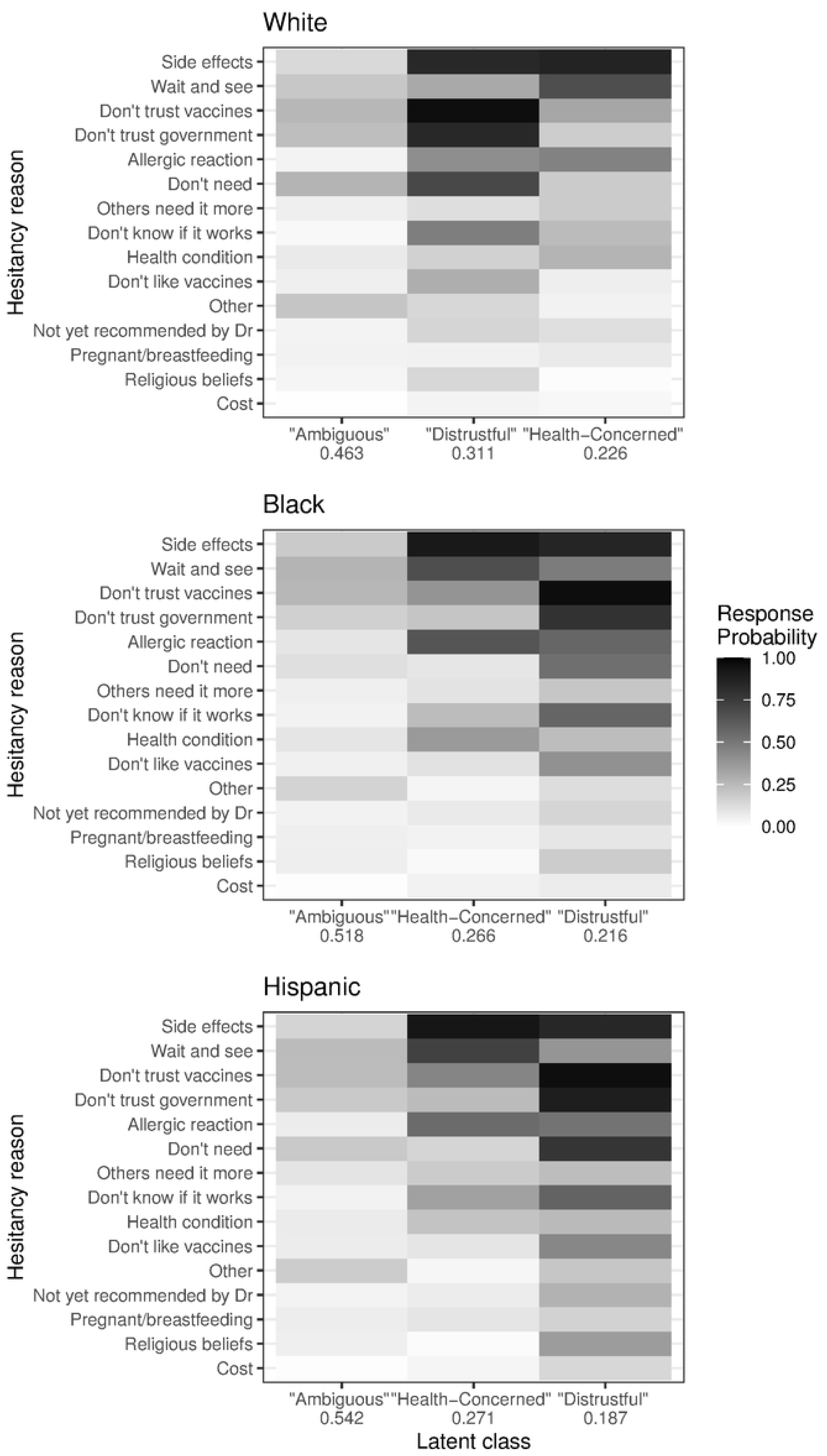
Results of LCA with three classes modeled on White, Black, and Hispanic respondents in May 2021. Hesitancy reasons are arranged according to overall prevalence (regardless of class or race/ethnicity) in the February sample, to facilitate comparison to the previous models.

In May, similar classes again emerged within each race/ethnicity sub-analysis: one ambiguous group characterized by low probabilities of individual selections, one dominated by distrustful reasons, and one focused on health concerns. Within each of the racial/ethnic groups, respondents are most likely to fall into the ambiguous category, but the relative size of this class decreased compared to February.

However, the models for each racial/ethnic group differ in the relative size of the second and third classes. As in the overall model for May 2021, White respondents had a higher probability of falling into the distrustful class (membership probability of 31.1%) and a lower probability for the health-concerned class (membership probability of 22.6%). However, for Black and Hispanic respondents, the health-concerned class remained more common (membership probabilities of 26.6% and 27.1%, respectively), compared to the distrustful class (membership probability of 21.6% for the Black group and 18.7% for the Hispanic group), mirroring the February results.

Also similar to the February analysis, the distrustful class for Black and Hispanic respondents differed from the distrustful class for Whites, again with Black and Hispanic respondents having a higher probability of selecting health-related reasons.

### Sensitivity Analysis

As noted above, the LCA model did not incorporate survey weights in the analyses. However, a sensitivity analysis (described in the Methods) found that the distribution of survey weights was similar across classes, indicating that inclusion of the weights would be unlikely to change the relative class size.

## Discussion

Latent class models fit on data from February and May 2021 revealed three distinct classes of COVID-19 vaccine hesitant respondents. Notably, similar classes arose when separate models were fit on the entire hesitant samples for February and May, and again, when separate models were fit on White, Black, and Hispanic subgroups at each of those time points. These results indicate that it is possible to identify consistent sets of belief patterns in all groups that were studied, both early in the vaccine rollout, when most people were not eligible for vaccination and thus reporting prospective hesitancy, and in May 2021, when all US adults were eligible and vaccine availability was widespread. Further, these belief patterns differ from each other in meaningful ways.

The two most interpretable classes identified by the model differ considerably. Health-concerned hesitancy emphasized concerns about side-effects, a desire to wait and see if the vaccine is safe, and concerns about allergies or not knowing if the vaccine works. Distrustful hesitancy was associated with a high probability of not trusting the COVID-19 vaccine, having concerns about side effects, and not trusting the government.

This analysis also highlights the challenges of studying vaccine hesitancy before a vaccine is available. Traditional vaccine hesitancy has been defined in terms of vaccination behaviors, such as “delay in acceptance or refusal of vaccination despite availability of vaccination services,” and factors including convenience, confidence, and complacency have been shown to play a role [4]. In this study, we define hesitancy as stated unwillingness to accept a vaccine were if offered to the person today. Conversely, vaccine acceptance was defined as a stated willingness to receive a vaccine, definition that included people who are not yet vaccinated among the vaccine accepting, even in May when eligibility was not an issue. This definition is similar to others developed to measure hesitancy to COVID-19 vaccines before they were widely available [5], but the lack of a clear measurement norm during this period highlights the challenge of measuring and understanding hesitancy to a new vaccine in during a public health emergency.

Despite those challenges, our findings suggest that some of the previously studied factors in vaccine hesitancy also play a role in COVID-19 vaccine hesitancy. For example, the emergence of the health-concerned class supports a need to build confidence in the COVID-19 vaccine, in line with previously studied dimensions of vaccine hesitancy. The emergence of a distrustful class could be viewed similarly, but is concerning, as it indicates reasons for COVID-19 vaccine hesitancy related to a larger distrust of government institutions, in line with research that has identified political environment and political views as predictors of COVID-19 [25]. Building confidence in government and institutions is likely related to political views, and a substantively different task from addressing health concerns.

An examination of the differences between February 2021, when the survey largely measured prospective hestitancy and May 2021, the point of universal vaccine eligibility, provides additional insights. Overall, the relative size of the hesitant population decreased, from 19.1% in February to 13.3% in May, in line with previous research (e.g., [12]). However, in May, 22% of unvaccinated respondents indicated that they definitely or probably would take a vaccine if it were offered to them that day, suggesting that access to the vaccine in May 2021 remained an issue.

Similar class types emerged across the two time points; however, their relative sizes in the sample changed. From February to May, the proportion in the ambiguous class decreased slightly and the relative size of the distrustful class increased from 22.1% to 27.6%, displacing health-concerned hesitancy as the second largest class. The decrease in the prevalence of hesitant respondents suggests that formerly hesitant respondents became vaccine accepting by May. This analysis cannot determine from which hesitant class those defections occurred, as the study did not track individual respondents over time. However, if hesitancy type persisted over the course of the rollout (that is, if the reason someone was hesitant in February did not change in May) and the primary change was conversion from vaccine hesitant to vaccine accepting, the change in relative class size would indicate indicate that the distrustful class was less receptive to efforts to increase vaccine acceptance.

Importantly, when we examined subgroups of White, Black, and Hispanic respondents, the flip of distrustful and hesitant class sizes was only seen for Whites. For Black and Hispanic participants, the health-concerned class was still larger than the distrustful class. If individuals who expressed health concerns in February were more open to being converted to vaccine accepting by May, these results support concerns about differential access to the vaccine during the period of limited supply. Previous research has shown that COVID-19 vaccine uptake between December 2020 and January 2021 varied by racial and ethnic groups [26] and was mediated by factors that varied between racial and ethnic groups, including employment, household income, marital status, and chronic health conditions, which contributed to lower rates of vaccination among Black and Hispanic populations compared to Whites [27, 28].

Also, distrust of the health care system among Black and Hispanic individuals in the US has been studied in general and in the context of COVID-19 vaccines [29], and vaccine hesitancy was expected among these populations. However, White hesitancy initially appeared lower [30] and was not as well studied. This study identified a potentially important distinction. At both time points, in the distrustful class for Black and Hispanic groups, health-related reasons had a higher probability of selection than in the White distrustful group. For example, “distrustful” Black and Hispanic respondents were more likely to have concerns about allergic reactions than an individual in the White distrustful class (16 and 11 percentage points, respectively) (see S1 Appendix). This suggests that for Black or Hispanic respondents, distrust of the vaccine or government was rooted in different concerns than the kind of distrust of the government that was prevalent among Whites in February and May, 2021, when the pandemic had been heavily politicized in the US. Research has shown that people living in counties with higher support from Trump in the 2020 presidential election were more likely to be vaccine hesitant [26], and those who identified as Democrats showed higher rates of vaccine acceptance than those who identified as Republican or Independent [25].

Additionally, during the vaccine rollout, outreach campaigns were initiated by many individuals and groups within their own communities, such as efforts by Black doctors and nurses [31]. Health organizations, including the CDC, provided resources and toolkits to promote vaccine uptake among non-White race and ethnic groups [32]. This study is not designed to evaluate specific interventions, but our results are consistent with the possibility that that the efforts of health providers, public health officials, and community leaders were to address the concerns of Black and Hispanic individuals were effective.

This study focused on the period of the initial COVID-19 vaccine rollout. However, the consistent emergency of the different hesitancy types suggests the identification of constructs with continued salience, as researchers continue to identify sources of continuing and increasing vaccine hesitancy [33].

## Limitations and Strengths

As the models used for this analysis do not support survey weights, the analysis is based on an unweighted sample recruited from the Facebook Monthly Active User Base, which is more female, White, and college educated that the US adult population. It was also not possible to link respondents across time to determine if respondents were included in both time periods. A sensitivity analysis to see if the relative size of classes would have been likely to changed if survey weights suggested such a change would be unlikely. Due to the scale of the survey, no fill-in option was available for the respondents to provide their own reasons for hesitancy, as the scale of the survey prohibited coding of write-in data, so responses were limited to the list of options provided on the survey. Additionally, this study relied on a common practice to categorize a Likert scale question on vaccine hesitancy into hesitant/not hesitant. Given the different hesitancy classes identified in this study, future work should examine whether those who say they “probably would not” or “definitely would not” get the vaccine show different hesitancy classes, or as seen in this study, different relative sizes of similar hesitancy classes.

A strength of the analysis is that the scale of the survey enabled important subgroup analyses, even when restricting the sample to only hesitant responses. Latent class analysis is only feasible for reasonably large data sets, which were available for this study. Additionally, the ability to compare responses from February and May enables insight into changes that occurred over the rollout, including responses public health campaigns, increased eligibility, and the possibility for individuals to see others receive a vaccine.

## Conclusion

COVID-19 vaccine hesitancy is a heterogeneous construct, and officials need to consider multiple, distinct types of hesitancy related to health concerns and government distrust. Further, in the US, types and their prevalence vary among racial and ethnic subgroups.

## Supporting information

**S1 Appendix Survey questions and response probabilities.** Full text of survey questions used in the analysis and tables of response probability estimates from each latent class model.

## Data Availability

Survey microdata is not publicly available because survey participants only consented to public disclosure of aggregate data. The microdata is available to researchers under a nondisclosure agreement permitting research uses. Access can be requested at https://www.icpsr.umich.edu/web/ICPSR/studies/39207

